# High prevalence of food insecurity, the adverse impact of COVID-19 in Brazilian favela

**DOI:** 10.1101/2020.07.31.20166157

**Authors:** Catarina Vezetiv Manfrinato, Aluízio Marino, Vitória Ferreira Condé, Maria do Carmo Pinho Franco, Elke Stedefeldt, Luciana Yuki Tomita

## Abstract

**Objective:** To investigate food insecurity prevalence in two favelas in Brazil in the early weeks from physical distancing policy, between March 27, 2020 to June 1, 2020.

**Design:** A cross-sectional study using online questionnaire to elicit information on socioeconomic and demographic characteristics, the types of stores visited to buy food and food insecurity screening. Experience of food insecurity was collected according to the Brazilian Food Insecurity Scale. Factors associated with moderate or severe food insecurity (EBIA≥3) were investigated using the logistic regression model.

**Setting:** São Paulo city, Brazil.

**Participants:** 909 householders.

**Results:** 88% of the households included young women working as cleaners or kitchen assistants and in sales services. One-fifth of the participants were recieving federal cash transfer programme, called Bolsa Família. There were 92% households with children. The most frequent experience reported was uncertainty about food acquisition or receiving more (89%), to eat less than one should (64%), not being able to eat healthy and nutritious food (46%), and skipping a meal (39%). 47% of the participants experienced moderate or severe food insecurity. Factors associated with moderate and severe food insecurity were low income, being Bolsa Família recipient, a low level of education, and households without children.

**Conclusions:** Half of the participants experienced moderate or severe food insecurity, and close to ten per cent was hungry. Our data suggest that families with children were at lower risk of moderate to severe food insecurity. It is possible that nationally established social programs like Bolsa Família were protecting those families.

## Introduction

The coronavirus disease 2019 (COVID-19) pandemic has led to the tragic loss of human life with deep social and economic consequences, including on food insecurity and nutrition. Disproportionate burdens of infections, hospitalisations, and deaths from COVID-19 among already vulnerable communities were observed^(1)^. Inequities in food and health access exacerbate inequalities in nutrition outcomes in the form of undernutrition and overweight individuals, obesity, and diet-related chronic diseases^(2)^.

The lack of regular access to nutritious and sufficient food experienced by such people puts them at greater risk of malnutrition, hidden hunger, or micronutrient deficiencies. The ability to eat a healthy diet is determined by the person’s access to affordable healthy food, the environment in which they live, and knowledge about the importance of those food groups for health^(3)^. People experiencing food insecurity may live in food deserts and may predominantly have access to low-cost, energy-dense processed foods^(3)^. Those calorie-dense processed foods are risk factors for obesity and obesity-related chronic diseases, such as diabetes, hypertension, and cardiovascular disease, and are strongly associated with severe COVID-19 outcomes. Obesity, hidden hunger, and coronavirus could face the triple burden of disease^(4)^.

In the COVID-19 pandemic, physical distancing or quarantine is impossible in regions such as the Brazilian favelas that house approximately 13 million individuals, which is crowded and has limited access to clean water or hygiene supplies^(5)^. Those people work in informal or less flexible jobs and are at a higher risk of losing their jobs completely or partially.

On 25 February 2020, the first COVID-19 case was confirmed in São Paulo city, Brazil. Twenty days later, community transmission was announced. One month after the first COVID-19 case, a physical distancing policy was adopted with the closing of schools and non-essential services.

The objective of the present study was to investigate food insecurity prevalence in two favelas in São Paulo city in the early weeks of the physical distancing policy.

## Methods

A cross-sectional study was conducted in two favelas in São Paulo city, Brazil, between 27 March and 1 June 2020. The inclusion criteria were those living in those communities and householders.

Heliopolis Favela is the biggest and most densely populated shantytown located close to the downtown area of São Paulo city. The population size is not precise. The last Brazilian census in 2010 estimated that 65 thousand people live in an area of approximately 1.2 km^2^, but local public health services indicate that they attend 140 thousand patients; the local non-profit organisation, the UNAS, estimated that 220 thousand people live there. Vila São José is a favela located in the south of São Paulo city. A projection of the local population from the Public Health Service is that 203 thousand people live in a 134 km^2^ area. On 24 May 2020, newspapers published reports that the peripheral regions of the city of São Paulo are where most people died from COVID-19 ^(6)^. The Vila São José community is located in a peripheral region.

### Data collection

A standardised online questionnaire was employed to elicit information on socioeconomic and demographic characteristics, the frequency of food purchases, the types of stores visited to buy food, finances, and food insecurity screening after the physical distancing policy came into effect. Data on the experience of food insecurity were collected according to the Brazilian Food Insecurity Scale^(7)^

An online questionnaire was collected in Heliopolis Favela by the observatory ‘De Olho na Quebrada’ (Eyes in the Broken). Each questionnaire was available for one week.

The household level of food insecurity was assessed using the short version of the Brazilian Food Insecurity Scale (EBIA), adapted from the US Household Food Security Survey Measure (HPSSM) and validated for the Brazilian population^(7)^. The short version is comprised of five questions and presents high sensibility and specificity compared to the original food insecurity scale^(8)^. The householder should answer the scale. A family that reported any experience of food insecurity, by answering “yes”, was scored as 1 point and 0 points for “no”, with a maximum of five points in the questionnaire. The five questions were based on assessing the perception or experience of food intake in the household since the beginning of social distancing: 1) anxiety and worry about the ability to obtain food; 2) too poor to buy more food; 3) whether the quality and variety of food has been compromised, including nutritious food; 4) quantity reduction and 5) skipping meals^(7)^. The total score classifications of the food insecurity grade were: (0) no insecurity, (1–2) mild insecurity, (3–4) moderate insecurity, and (5) severe insecurity.

The income per person was converted from Brazilian Real to U.S. Dollars using the currency conversion rate during the period.

Ethical approval was obtained from the Institutional Review Board of the Universidade Federal de São Paulo and the Medical Ethical Committees of the participating hospitals (CAAE 30805520.7.0000.5505). Online consent was obtained from all participants. The present study complies with the Strengthening the Reporting of Observational Studies in Epidemiology guidelines.

### Statistical analysis

Crude and relative distributions and median and interquartile ranges were calculated for the descriptive statistics.

Factors associated with moderate or severe food insecurity (EBIA≥3) were investigated using the logistic regression model. The reference category for exposure of interest was the lowest risk for food insecurity. For income, the first quartile among households with no food insecurity was the reference category. A level of significance of 5% was adopted.

Statistical analysis was performed using STATA 14.0. (Texas, USA).

## Results

In the present study, 1 172 participants answered the online survey. Of these, 123 (10%) did not live in those communities, 86 (7.3%) were not householders, and 54 (4.6%) answers were repeated. In Heliopolis, a questionnaire was administered twice. The first was administered on 27 March, four days after the social distancing policy came into effect, collecting 653 responses. The second was administered after one month, on 30 April, at which time 756 participants assessed their experience of food insecurity. For the present study, only participants who answered both questionnaires were considered. Of these, 909 (78%) were considered eligible participants; 697 lived in Heliopolis Favela and 212 in Vila São José.

The majority of the households included young women working as cleaners or kitchen assistants and in sales services. One-fifth were Bolsa Família recipients, a federal cash transfer programme for families with children that attend school. There were many overcrowded households with children.

The majority of participants purchased food in a local supermarket (55%) or a local market close to home (43%), and only 2% reported buying food in *feira*, an outdoor public market that sells fresh vegetables and fruits; 98% observed an increase in food prices. Foods were purchased monthly (66%), twice a month (23%), and weekly (8%).

More than half of the participants were in moderate and severe food insecurity (56%). The most frequent experience reported was uncertainty about food acquisition or receiving more, to eat less than one should, not being able to eat healthy and nutritious food, and skipping a meal. A quarter reported that food was consumed before buying or receiving more (Table 1).

**Table 1.** Brazilian’s favela household characteristics and COVID-19 impact after social distancing, Brazil, April, 2020 (n=909)

| **Brazilian's favela** | **n (%) or P50 (P25, P75)** |
| --- | --- |
| **Householder characteristics** |  |
| Female (%) | 796 (88%) |
| Age, years | 33 (28, 39) |
| Monthly per capita income, USD | 70.2 (46.6) |
| Bolsa família recipients | 236 (26%) |
| Occupation |  |
| Cleaner, kitchen assistant, driver, deliveryman | 369 (40%) |
| Salesman, freelance, autonomous | 290 (33%) |
| Unemployed, housewife, retired | 211 (23%) |
| Graduated | 39 (4%) |
| **Household characteristics** |  |
| Number of dwellers | 4 (3,5) |
| Household with child under 10y | 836 (92%) |
| **Food insecurity** |  |
| Score at risk of food insecurity* | 3 (2, 4) |
| No insecurity | 47 (5%) |
| Mild insecurity | 353 (39%) |
| Moderate insecurity | 426 (47%) |
| Severe insecurity | 83 (9%) |
| Uncertainty about food acquisition or receiving more | 807 (89%) |
| Ran out of food before buying or receiving more | 208 (23%) |
| Unable to eat healthy and nutritious food | 421 (46%) |
| Ate less than one should | 584 (64%) |
| Skipped a meal | 350 (39%) |

\* no insecurity (0), mild insecurity (1-2 points), moderate insecurity (3-4 points), severe insecurity (5points)

Factors associated with moderate and severe food insecurity were low income, being a Bolsa Família recipient, a low level of education, and households without children (Table 2).

**Table 2.** Associated factors for moderate and severe insecurity after social distancing from COVID-19, Brazil, April, 2020

| Characteristics | No food insecurity or mild food insecurity | Moderate or severe food insecurity | OR (95%CI) |
| --- | --- | --- | --- |
|  | n(%) | n(%) |  |
| Income per person, USD |  |  |  |
| $\geq 62$ | 223 (62) | 200 (48) | 1.00 |
| $\leq 61$ | 134 (38) | 217 (52) | 1.81 (1.35-2.41) |
| Bolsa família recipient* |  |  |  |
| no | 309 (77) | 364 (72) | 1.00 |
| yes | 91 (23) | 145 (28) | 1.35 (1.00-1.83) |
| Years of education |  |  |  |
| $\geq 9$ years | 221 (56) | 203 (40) | 1.00 |
| $\leq 8$ years | 176(44) | 303 (60) | 1.87 (1.44-2.44) |
| Household with children |  |  |  |
| yes | 383 (96) | 453 (89) | 1.00 |
| no | 17 (4) | 56 (11) | 2.77 (1.59-4.76) |

\* federal cash transfer program

## Discussion

Our study presents the impact on food access immediately following the COVID-19-related physical distancing measure and school closure in two favelas in São Paulo city, Brazil. More than half of the participants experienced moderate or severe food insecurity, and close to ten per cent were hungry. Half were unable to eat healthy and nutritious food, while families with children were at a lower risk for insecurity. Almost all participants reported that food prices increased and the majority purchased food from supermarkets, but not from local outdoor markets where fresh vegetables and fruits are available.

Unfortunately, a downward trend in food insecurity observed in Brazil from 2004 to 2013, 17% to 7.9% respectively, has been diverted^(9)^. At that time, the associated factors of moderate and severe food insecurity were living in the Northeast and Northern regions, being among the poorest and living in an urban area with inadequate sanitation, a household density of more than two persons per bedroom, less than four household material goods, and households headed by females, individuals younger than 60, non-whites, having less than four years of schooling, and being unemployed. These associated factors remained in the ten years covered by the National Survey^(9)^.

Early effects of the COVID-19 pandemic were also observed among low-income Americans, where 44% of households were food insecure^(10)^. However, among Americans, an increase in household food insecurity was observed in previous years, from 11% in 2018 to 38% in March 2020. People who were non-Hispanic, Black, or Hispanic, had children in the home, and had less than a college education were more likely to be at risk of food insecurity. According to the United Nations Department of Economic and Social Affairs (2020), the COVID-19 pandemic has affected all segments of the population, but it was reinforced among the poorest and most vulnerable people. Many of these people are workers in the informal economy, as observed in our study^(11)^.

In the Heliopolis Observatory, collected in the first week after COVID-19 physical distancing measures were enacted, 65% reported that they were not working due to social distancing, 68% reported considerable income reduction, and 17% reported no wages, with higher impacts occurring among the poorest people^(12)^. A Brazilian Federal cash transfer of USD 110 was offered for informal workers after a social appeal, one month after the social distancing policy came into effect. However, according to the observatory, among the 83% who asked for emergency cash, only one-third received it.

As a result of the economic impact of COVID-19, the number of people around the world facing acute food insecurity is 265 million in 2020, an increase of 130 million from 135 million in 2019 according to the World Food Programme projection (WFP)^(11)^. The WFP recommended measures, including pre-positioning food closest to those most in need while supply chains are still working, providing double food rations per person to reduce the number of distributions, providing take-home food rations to replace school meals, and launching health-education campaigns^(12)^. For many students around the world, school feeding is the only meal they receive during the day. To protect students from food insecurity, the São Paulo state government offered an additional cash payment of USD 10 per month for Bolsa Família recipients for 27% of students. However, in our study, only 15% received it^(12)^. The distribution of food kits, called *cesta básica* in Portuguese, from the São Paulo city prefecture, comprised of 25 kg minimally processed food such as rice and beans, processed foods such as pasta, salt, sugar, tomato pasta, green beans, corn, maize flour, and vegetable oil, as well as ultra-processed food such as biscuits. This food kit was also offered to student recipients of Bolsa Família.

The Brazilian public agenda has an intersectorial and participatory approach called Food and Nutrition Security, that aims to develop public policies to guaranty food and nutrition security. This concept is to ensure human rights for a healthy, accessible, and adequate diet, without compromising access to other essential needs, such as respecting healthy eating practises and cultural diversity, and should be socioeconomically and agro-ecologically sustainable^(13)^. Through this program, family farming from settlements should be supported in terms of sales to benefit socially vulnerable and marginalised populations, such as quilombolas (Afro-Brazilian ancestry), indigenous people, and for school feeding, entitled Programa de Aquisição de Alimentos (PAA) or Program for Food Acquisition in English^(14)^. In the present situation of poor access to healthy food in a highly vulnerable community, this program should work. However, in the actual government, financing for social programs has been reduced, including the PAA. Over the past fifteen years, the PAA has shown how a Brazilian public policy has importance in boosting local economies and short production/distribution circuits. In addition, the PAA also supports the structuring role and income provision for family farming and the guarantee of the human right to adequate food ^(15)^. It would be essential in the context of the pandemic to offer logistical support so that farmers can directly market their products to consumers in urban centres, reducing the risk of COVID-19. At the same time, safe conditions must be offered for production to be acquired by the government for the distribution of food baskets. The strategic use of PAA in the crisis would also involve reactivating the modalities of direct purchases and stock formation. Moreover, the program’s institutional channels can serve federal resources and be transferred to states and municipalities to finance local strategies for responding to the effects of the pandemic ^(15)^.

Our data suggest that families with children were at lower risk of moderate to severe food insecurity. It is possible that nationally established social programs like Bolsa Família were protecting those families, as well as solidarity from non-profit organisations and private sectors that offer *cesta básica* instead of government defaults during the COVID-19 pandemic period.

A systematic review observed that female-headed households were 75% more likely to be food insecure than male-headed households. This could be because of the lower income earned by women compared to men due to the sex wage difference or reduced work time among women, the greater observation of household needs and food preparation among women than men^(16)^ and the increased childcare burden shouldered by female household providers, such as feeding and education.

The impact of COVID-19 among Brazilian families could be significant. Brazil was already experiencing food insecurity before the coronavirus pandemic: more than half of adults were excess weight and 19.8% were obese, 24.7% were diagnosed with hypertension, and 7.7% with diabetes, with a higher prevalence of morbidity among those with low levels of education; only 23% reported an intake of five or more portions of vegetables and fruits, with a lower frequency observed among those with lower levels of education^(17)^. Among adolescents, a cross-sectional national study showed that 3.3% were overweight, 21.3% were obese, and a premature impact on their health was observed: 20% presented hypercholesterolaemia, 47% low HDL cholesterol, 7.8% hypertriglyceridaemia, 8% hypertension, 4% had high glucose levels, and 2.6% suffered from metabolic syndrome^(18,19)^. In the two editions of the National Adolescent School-based Health Survey, 2009 and 2012, the consumption of beans and fruits decreased; in the most recent survey, 60% reported bean intake and 29.8% fruits, 35.4% sweet beverages, and 42.6% reported to consuming sweets at least five days a week^(20)^. Together, the antagonistic health-related problems and double burden of malnutrition among Brazilian adolescents with a high body mass index (BMI) and low height for their age was observed at 0.3%^(21)^.

The quality of food that was stocked up on before social distancing came into effect is a concern, as almost all participants reported buying food in supermarkets and only two per cent reported doing so in outdoor fresh vegetable and fruit stores. The increased consumption of ultra-processed food combined with reduced physical activity during the pandemic period could worsen or increase the number of overweight people and chronic diseases, risk factors that have been related to increasing the severity of COVID-19. An estimate obtained in a nationally representative Brazilian household-based health survey pointed out that 34% of adults present at least one risk factor for severe COVID-19^(22)^. In São Paulo city, the prevalence of one or more risk factors for severe COVID-19 was 56%, with a higher prevalence among less educated adults (86%) compared to those with a university education (49%); a similar distribution was seen according to income or race^(23)^.

The limitations of the present study should be considered. It is a cross-sectional study, and no temporality and causality can be inferred. No statistical probabilistic sample was drawn since the population size of the communities in question is unknown. The web questionnaire was shared between both communities through local acting non-profit organisations and community leaders. It is possible that it reached many households due to their high capillarity and relevance to social policy. However, families that are going hungry could be under-represented since they may not have an internet plan or mobile phone.

The present study showed the importance and swift impact of social distancing in São Paulo’s favelas. These findings should be considered when designing and implementing social policy intended to act fast, closing family farming to socially vulnerable communities, as well as implementing assistance programs, especially for households without children, who are more likely to go hungry.

## Data Availability

None data available.

## Acknowledgments

We thank members from the Observatory “De Olho na Quebrada”, André Luis Silva, Gabriel Feitosa, Gabrielle Souza, João Victor da Cruz, Karoline Aparecida, Leonardo da Silva Pimentel, Letícia Avelino, Edgard Barki, Marina Lima, Isabela Lemos, Reginaldo José Gonçalves and Vila São José’s community leader Luiz Alberto F. Alves and CAPES (Coordenação de Aperfeiçoamento de Pessoal de Nível Superior) for CVZ scholarship and financial code 001.

